# Sensitivity and specificity of a rapid test for assessment of exposure to SARS-CoV-2 in a community-based setting in Brazil

**DOI:** 10.1101/2020.05.06.20093476

**Authors:** Lucia Campos Pellanda, Eliana Márcia da Ros Wendland, Alan John Alexander McBride, Luciana Tovo-Rodrigues, Marcos Roberto Alves Ferreira, Odir Antônio Dellagostin, Mariangela Freitas da Silveira, Aluisio Jardim Dornellas de Barros, Pedro Curi Hallal, Cesar Gomes Victora

**Author notes:** Corresponding author: Lucia C Pellanda, Universidade Federal de Ciências da Saúde de Porto Alegre, Rua Sarmento Leite, 245 - Porto Alegre, Rio Grande do Sul, Brazil - CEP 90050-170, Fone.

## Abstract

**Background:** While the recommended laboratory diagnosis of COVID-19 is a molecular based assay, population-based studies to determine the prevalence of COVID-19 usually use serological assays.

**Objective:** To evaluate the sensitivity and specificity of a rapid diagnostic test for COVID-19 compared to quantitative reverse transcription polymerase chain reaction (qRT-PCR).

**Methods:** We evaluated the sensitivity using a panel of finger prick blood samples from participants >18 years of age that had been tested for COVID-19 by qRT-PCR. For assessing specificity, we used serum samples from the 1982 Pelotas (Brazil) Birth Cohort participants collected in 2012 with no exposure to SARS-CoV-2.

**Results:** The sensitivity of the test was 77.1% (95% CI 66.6 - 85.6), based upon 83 subjects who had tested positive for qRT-PCR at least 10 days before the rapid diagnostic test (RDT). Based upon 100 sera samples, specificity was 98.0% (95% CI 92.9 - 99.8). There was substantial agreement (Kappa score 0.76) between the qRT-PCR results and the RDT.

**Interpretation:** The validation results are well in line with previous assessments of the test, and confirm that it is sufficiently precise for epidemiological studies aimed at monitoring levels and trends of the COVID-19 pandemic.

## Introduction

The COVID-19 pandemic continues to cause havoc around the world, and as of May 2nd there were 3,2362,772 confirmed cases and 239,227 fatalities reported [1]. Due to supply chain problems, many governments have opted to test only the most serious cases such that the epidemiology of the disease remains largely unknown. This has led public health experts to believe that the prevalence of COVID-19 is significantly underestimated [2,3].

The laboratory reference method for detecting COVID-19 is based on real-time reverse transcriptase PCR (qRT-PCR), to detect the presence of SARS-CoV-2 in nasofaringeal swabs collected from patients suspected to be infected [4]. While it is highly specific, there are reports of low sensitivity in the field [3,5–7]. The ability of the qRT-PCR to detect SARS-CoV-2 is dependent on the presence of the virus in the sample and this is subject to several variables, e.g. the virus must be present in the patients nasopharinx at the time of collection and the correct use of the swab [8]. The failure to meet the necessary criteria for collection can potentially result in a false negative result. This has dire consequences for public health, especially for asymptomatic individuals as they will continue to infect others as they are not aware of the risk of transmission [7], leading to the need of evaluating other diagnostic tools that could be useful in these settings.

Public health services around the world have resorted to purchasing serological assays towards overcoming the lack of availability of the standard nucleic acid tests. Given the urgent need for diagnostic tests, many regulatory agencies approved the emergency use of diagnostics based on the manufacturer’s reports of diagnostic accuracy without the need for further validation [9,10]. Serological tests identify antibodies in symptomatic and asymptomatic individuals and, although of limited clinical use in diagnosis infection [11], are important in population-based studies that are necessary to estimate the seroprevalence of COVID-19. However, given the current global emergency, very few studies have evaluated the diagnostic properties of rapid diagnostic tests (RDT) in the settings where they will be used. Furthermore, there have been reports in the literature of significant discrepancies between the manufacturer’s performance claims and the results from the field [4,9].

The time required for the immune system to produce detectable levels of immunoglobulins typically ranges from 5 to 10 days, but this may take longer in the case of SARS-CoV-2 [7,12,13]. The Immunochromatographic Lateral Flow Assay (Wondfo SARS-CoV-2 Antibody Test, China)[14] used in this study detects IgG and IgM combined antibodies in blood, plasma or serum samples. This test was validated in a study performed by the manufacturer, which reported a sensitivity and specificity of 86.4 and 99.6%, respectively [14]. These tests were purchased in bulk by the Brazilian government and they were earmarked for use in population surveys and surveillance programs. An initial validation study was carried out by the National Institute for Quality Control in Health [15] using 18 qRT-PCR positive and 77 negative serum samples. The reported sensitivity was 100% (95% CI 81.50-100), while specificity was 98.7% (95% CI 92.98-99.97), performance characteristics similar to those claimed by the manufacturer. However, as the RDT was to be used in a large-scale, population-based household survey of COVID-19 in the state of Rio Grande do Sul in Southern Brazil, we decided it was necessary to evaluate the RDT with an expanded panel of sera collected from the target population.

## Methods

### Ethical approval

This study and the protocols used therein were approved by the National Commission on Ethics in Research (CONEP, Brasilia, DF). All participants signed an informed consent form.

### Study design

The study population included individuals (>18 yr. old) both symptomatic or asymptomatic at the time of testing and who were qRT-PCR positive for SARS-CoV-2 following the testing of nasopharyngeal swabs during March 2020. At least 10 days later (to allow for the production of anti-SARS-CoV-2 antibodies), the study participants were invited to complete a questionnaire on their COVID-19 symptoms and demographic information. A blood sample was collected by fingerstick and the RDT was carried out at the time of collection as recommended by the manufacturer.

Samples from healthy individuals (n = 100) were randomly selected from a pre-COVID-19 serum collection from 3,700 individuals belonging to the 1982 Birth Cohort Study [16] and stored at the Epidemiology Research Centre, Federal University of Pelotas (UFPel, Pelotas, RS).

### Sample preparation

Fingerstick blood samples were applied directly to the RDT. Frozen serum samples were thawed at room temperature, gently shaken, and centrifuged at 10,000 × g for 10 min at 4°C. A random subsample of 15 serum samples was tested by dot-blot for the presence of IgG, IgM and IgA, confirming sample integrity.

### RDTprocedure

The assay used in this study was the Wondfo SARS-CoV-2 Antibody Test (Lateral Flow Method). The assays were carried out according to the manufacturers protocol: a 10 μl sample (whole blood or serum) was applied to the sample well, followed by the addition of 2-3 drops or 80μl of diluent. The test was developed for 15 minutes at room temperature and the results (positive or negative) were read by independent experienced readers blinded to the sample status. Only RDTs whose control line was positive were included in the statistical analysis. Note, while the RDT can detect both IgM and IgG anti-SARS-CoV-2 antibodies, it does not discriminate between antibody types.

### Data analysis

Diagnostic accuracy was calculated using sensitivity and specificity, and agreement to the reference test (qRT-PCR) was assessed using Cohen’s Kappa. The 95% confidence intervals for all estimates were calculated using the exact method. Statistical analyses were performed using MedCalc for Windows, version 19.2 (MedCalc Software, Ostend, Belgium).

## Results

Volunteers were invited through newspaper and social media to participate in the RDT study; of the 95 qRT-PCR positive individuals who responded, 83 had been diagnosed 10 or more days previously and were enrolled in the study. The majority of the qRT-PCR positive participants were man (53.0%), self-reported white skin color (96.0%), and 18 (21.7%) had at least one symptom associated with COVID-19 when tested with the RDT. The mean age for the group was 48.6 (SD = 14.4) years. In what regards the pre-COVID-19 group, 37.0% were man, 82.0% self-reported white skin color. The mean age for the group was 30.0 (SD = 0.3) years.

The sensitivity of the RDT among those previously diagnosed with COVID-19 was 77.1% (64/83) and the specificity was 98.0% in the pre-COVID-19 healthy individuals, see Table 1. When compared to the reference method, the RDT demonstrated substantial agreement with the qRT-PCR. As the prevalence rate of COVID-19 remains unknown, we calculated the predictive values of the RDT based on prevalence rates of 2, 4 and 10%; the PPVs ranged from 44 to 81% and the NPVs varied between 97.5 and 99.5%.

**Table 1.** Performance characteristics of the COVID-19 RDT.

| % Sensitivity<br>(95% CI) | % Specificity<br>(95% CI) | Kappa Agreement<br>(95% CI) | % PPV<br>(95% CI) | % NPV<br>(95% CI) |
| --- | --- | --- | --- | --- |
| 77.1 (66.6 - 85.6) | 98.0 (92.9 - 99.8) | 0.76 (0.67 - 0.86) | 44.0 (16.5 - 75.7)<sup>1</sup> | 99.5 (99.3 - 99.7)<sup>1</sup> |
|  |  |  | 61.6 (28.8 - 86.4)<sup>2</sup> | 99.0 (98.6 - 99.4)<sup>2</sup> |
|  |  |  | 81.1 (51.9 - 94.4)<sup>3</sup> | 97.5 (96.3 - 98.3)<sup>3</sup> |

<sup>1</sup>Prevalence rate = 2%; <sup>2</sup>4%; <sup>3</sup>10%. CI – confidence interval; PPV – positive predictive value; NPV – negative predictive value.

## Discussion

This study reports the evaluation of an RDT using samples from confirmed COVID-19 infected individuals and a pre-COVID-19 collection from healthy individuals. While the recommended laboratory test for COVID-19 is based on qRT-PCR assay, serological assays have an important role to play in epidemiological studies of the COVID-19 pandemic. The qRT-PCR has good performance characteristics [8,15]. In controlled laboratory conditions the sensitivity and specificity were 100% and 100%, respectively [10]. Yet, it is a complex technique and there are reports that it has been difficult to implement and results from the field have been disappointing [3,5-7], with sensitivity as low as 52% [8]. In addition, in Brazil there are significant problems in the supply of the kit to state central diagnostic laboratories. These shortages mean that only the most serious patients are being tested and this has made it difficult to draw any meaningful conclusions as to the transmission, prevalence and mortality rates of COVID-19.

Initial reports suggest that following infection with SARS-CoV-2, the immune system takes 6-21 days to product IgM and IgG antibodies [5,7,17,18]. Indeed, the RDT manufacturer states that antibodies only become detectable 7 days after disease onset. This observation was factored into the inclusion criteria used in the current study; samples were only collected from volunteers whose qRT-PCR test had been carried out at least 10 days previously.

The manufacturer of the RDT reported a sensitivity of 86.4% and a specificity of 99.6% using samples collected from 596 individuals (361 confirmed cases and 235 negative controls). In the current study, the sensitivity of the RDT was determined to be 77.1% (95% CI 66.6 - 85.6) and the specificity was 98.0% (95% CI 92.9 - 99.8). Although the diagnostic accuracy of the current study was lower than the original study, this was not unexpected given the geographic and genetic differences between the study populations. Our results are in accordance with a validation study performed in the United States, in which the sensitivity ranged from 40% to 82% (5 and >20 days after the symptoms onset, respectively) while the specificity was 99.06% [19].

Some limitations of our study merit discussion. First, we included two different samples to study sensitivity (individuals with previous positive qRT-PCR) and specificity (sera from the Pelotas Cohort Study). Both samples are similar in percentage of self-reported skin color, but the specificity sample was younger. Also, only the second sample is population based. Although sensitivity and specificity are intrinsic diagnostic properties of the test and theoretically do not vary with prevalence, it is important to consider that our positive sample set may not be representative of the entire spectrum of disease.

However, it included both symptomatic and asymptomatic individuals at the time of testing, representing an important population regarding the need of testing.

This means that the test may have even higher accuracy in more serious disease.

The second limitation is inherent to the reference method. The reference method based on qRT-PCR to detect the presence of SARS-CoV-2 in nasofaringeal swabs collected from patients suspected to have COVID-19 is highly dependent on the quality and timing of sample collection [3,5,7]. When the reference test does not present with 100% specificity or sensitivity, true results of the study test may be interpreted as false. This may happen specially when the NAAT fails to detect disease, and qRT-PCR false negatives may result in misinterpretation of a RDT positive result.

It is also important to emphasize that NAATs and RDTs are designed to be used in different clinical settings, respectively, diagnosis of acute disease and population evaluation of contact with SARS-CoV-2. While the true prevalence rate of COVID-19 remains unknown, several studies based on seroprevalence among the tested individuals have suggested that COVID-19 prevalence varies from less than 1% to 5.6% [9,20,21]. Furthermore, there are reports that the prevalence rate could be as high as 21% [22]. At a prevalence rate of 2% the PPV was 44%, meaning that just under half of the individuals with a positive RDT result will have the disease and this increased to over 81% when the prevalence rate was 10%. These are important observations and demonstrate that this RDT is suitable for a populational study to determine the levels and trends in the prevalence of COVID-19, especially when there is no need for a precise diagnosis at individual level.

## Data Availability

Due to the sensitive nature of the questions asked in this study, survey respondents were assured raw data would remain confidential and would not be shared.
Data not available / The data that has been used is confidential

CRediT author statement

**Lucia C Pellanda:** Conceptualization, Methodology, Data curation and analysis, Writing-Original draft preparation and Editing.

**Eliana Márcia da Ros Wendland:** Data collection, curation and analysis; Writing-Reviewing and Editing.

**Alan J A McBride:** Conceptualization, Formal analysis, Writing-Original draft preparation and Editing

**Luciana Tovo-Rodrigues:** Data collection; Writing-Reviewing and Editing.

**Marcos Roberto Alves Ferreira:** Data collection, Formal analysis

**Odir A Dellagostin** Conceptualization, Writing-Reviewing and Editing,

**Mariangela Freitas Ferreira** Data curation, analysis, Writing-Reviewing and Editing,

**Aluísio J D Barros** Writing-Reviewing and Editing,

**Pedro Curi Hallal** Conceptualization, Methodology, Writing-Reviewing and Editing, project administration, funding acquisition

**Cesar Gomes Victora** Conceptualization, Methodology, Data curation, analysis, Writing-Reviewing and Editing,, funding acquisition Role of the funding source

This work started through the Data Committee created by the State of Rio Grande do Sul government to fight the COVID-19 pandemics. The tests used in the study have been provided by the Brazilian Ministry of Health. Financial support for for data collection was provided by UNIMED Porto Alegre, Instituto Cultural Floresta and Instituto Serrapilheira. None of these funding sources took any part in study conceptualization, data collection, analysis or manuscript preparation, reviewing, or in the decision to submit the article for publication.

## Acknowledgements

The authors thank Alessandra Dahmer, Ana Carolina de Moura, Ana Paula Palauro Goularte, Dinara Jacqueline Moura, Emerson Silveira de Brito, Indianara Franciele Porgere, Giovana Petracco de Miranda, Giovana Tavares, Giulia Souza, Luana Freese, Lara Goulart Garcia, Marilia Mesenburg, Michele Paula Pretto, Natan José Dutra Dias, Roberta Beux, Suelen Porto Basgalupp, Thayane Martins Dornelles, Tiago Fetzner and William Jones Dartora

## Data statement

Due to the sensitive nature of the questions asked in this study, survey respondents were assured raw data would remain confidential and would not be shared.

*Data not available / The data that has been used is confidential*

